# The reproduction number R for COVID-19 in England: Why hasn’t “lockdown” been more effective?

**DOI:** 10.1101/2020.07.02.20144840

**Authors:** Alastair Grant

## Abstract

The reproduction number R, the average number of people that a single individual with a contagious disease infects, is central to understanding the dynamics of the COVID-19 epidemic. Values greater than one correspond to increasing rates of infection, and values less than one indicate that control measures are being effective. Here, we summarise how changes in the behaviour of individuals alter the value of R. We also use matrix models that correctly recreate distributions of times that individuals spend incubating the disease and being infective to demonstrate the accuracy of a simple approximation to estimate R directly from time series of case numbers, hospital admissions or deaths. The largest uncertainty is that the generation time of the infection is not precisely known, but this challenge also affects most of the more complex methods of calculating R. We use this approximation to examine changes in R in response to the introduction of “lockdown” restrictions in England. This suggests that there was a substantial reduction in R *before* large scale compulsory restrictions on economic and social activity were imposed on 23^rd^ March 2020. From mid-April to mid-June decline of the epidemic at national and regional level has been relatively slow, despite these restrictions (R values clustered around 0.81). However, these estimates of R are consistent with the relatively high average numbers of close contacts reported by confirmed cases combined with directly measured attack rates via close interactions. This implies that a significant portion of transmission is occurring in workplaces; overcrowded housing or through close contacts that are not currently lawful, routes on which nationwide lockdown will have limited impact.

## Introduction

In many countries, the emergent COVID-19 epidemic has shown an initial phase of exponential growth (Huang, Yang, Dai, Tian, & Chen, 2020). Governments have responded to this with the introduction of a range of infection control measures, including extensive restriction of social and economic interactions, often referred to as “lockdown” (see Jarvis et al., 2020, for a more detailed discussion of the UK regulations). The effective reproduction number of the virus, R, is widely used as a measure of epidemic growth during the initial unrestricted growth phase, and as a measure of the effectiveness of infection control measures after these have been introduced. R > 1 indicates that case numbers are growing, while R < 1 indicates falling infection rates. A number of methods are available to estimate R from data, such as time series of infection rates, hospitalisation rates or deaths (e.g. Britton & Tomba, 2019; Chong, 2020; Cori, Ferguson, Fraser, & Cauchemez, 2013; Fraser, 2007; Ganyani et al., 2020; Government Office for Science, 2020; Wallinga & Lipsitch, 2007). Some studies take into account population age structure and age-specific mortality rates and in some cases aggregate multiple data sources including deaths, antibody prevalence and information on contacts between individuals (Birrell et al., 2020). A number, but not all, of these calculations rely on compartment models such as SEIR (Belfin, Brodka, Radhakrishnan, & Rejula, 2020; K. Prem et al., 2020; Ridenhour, Kowalik, & Shay, 2014; J. H. Wu, Tang, Bragazzi, Nah, & McCarthy, 2020), which track numbers of susceptible, exposed, infective and recovering individuals. However, compartment models are known to give relatively poor predictions of epidemic dynamics as they do not correctly simulate the distribution of times between initial infection and either onset of symptoms or death (Grant, 2020). Although the meaning, and importance, of R is clear, this reliance on complex mathematical models makes it is difficult to understand the relationships between R and readily observable measures of the course of the epidemic, such as time series of infection rates, hospitalisation rates or deaths (Ridenhour et al., 2014). Here we present a simple model of the dynamics of COVID-19 infection that overcomes the problems in using compartment models to understand epidemic dynamics. We use this to demonstrate the accuracy of a simple approximation that allows R to be directly estimated from time series, then apply this to data on death and infection rates in England. We also outline the links between the behaviour of individuals and changes in the value of R, allowing us to examine whether our current understanding of behavioural responses to advice and regulation are consistent with the observed reductions in R.

### How is the value of R linked to people’s behaviour?

For an infection that is passed directly from one individual to another, R integrates the outcomes of all interactions that an individual has during the period that they are infectious. R is, therefore, dependent upon the period of time that an individual is infectious; the number of other individuals encountered per day; the probability of the disease being transmitted during an encounter and the proportion of the population that is susceptible to infection.

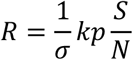

Where 1/σ is the duration of the infective period (expressed as a reciprocal to preserve standard notation used in compartment models); k is the average number of people encountered per day, p is the average probability of infection transmission at each encounter and S/N is the proportion of the population that is susceptible to infection. For a novel infective agent, the whole population is susceptible. S = N so S/N = 1, and R is then known as R_0_. This calculation is phrased in terms of average transmission and encounter rates, whereas there will be variations between individuals in how many other people they encounter each day (c.f. Mossong et al., 2008) and variation between encounters in their closeness and duration. For example, Bi et al. (2020) found that attack rates between household members were more than ten times higher than for contacts with others, and there was a 32 fold difference of attack rates depending upon whether close contacts between individuals were “rare” or “often”. The high rates of death in occupations such as security guards (ONS, 2020a) presumably reflects encounters with large numbers of people, with a rather low probability of transmission from each individual encounter. This formulation also assumes that the duration of the infective period and rates of transmission during this are constant, whereas many studies indicate that both of them vary (Britton & Tomba, 2019; Lauer et al., 2020). Transmission may be possible for some time before individuals become symptomatic (He et al., 2020), and symptomatic individuals may reduce their interactions with others through self-isolation. But the formula can be straightforwardly generalised to:

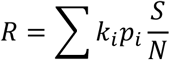

Where k_i_ and p_i_ are daily encounter and infectivity rates, and the summation is over the whole infective period. The daily infectivity can take into account both variation in viral shedding over time and variations in the duration of the infective period. This formulation clearly displays how the spread of an epidemic can be controlled by reducing numbers of contacts between individuals (bringing down the value of k), as well as by reducing the probability on transmission, p, during each encounter by measures such as hand washing, avoiding skin contact, maintaining larger social distances than previously and self-isolation by individuals who are symptomatic.

Estimates of R_0_ for COVID-19 mostly lie between 2 and 4, although some are above this range and in the confined conditions of the Diamond Princess cruise ship R_0_ may have been as high as 14.8 (Belfin et al., 2020; Rocklöv, Sjödin, & Wilder-Smith, 2020). If infectivity remains constant then reductions in R will be proportional to reductions in the average number of contacts between individuals. In a 2008 study across 8 European countries, individuals reported a mean of 13.4 contacts per day, but data were right skewed with some reporting more than 50 contacts a day (Mossong et al., 2008). In the UK, a survey using similar methods carried out after the introduction of a requirement for people to stay at home, except in very restricted circumstances, found that numbers of contacts were 74% lower than the UK data in the 2008 study (reducing from 10.8 to 2.8). Skin to skin contacts were reduced even more (Jarvis et al., 2020). The authors estimated that these would correspond to a reduction in R from 2.6 before social distancing to 0.62 based on all contacts and 0.37 based on transmission via skin contact alone. The changes in behaviour may also have altered the value of p, as the proportion of contacts that occurred at home increased from 34% to 58%. The average probability of transmission from an infective individual to a single contact will have increased, but only because contacts with those from outside of the home, which have a lower individual transmission probability, have reduced. This will lead to reduced transmission *between* homes and, in time, mean that many of the contacts of infective individuals will be with the previously infected (and therefore immune) individuals in the same household from who they acquired the infection. In Wuhan and Shanghai, mean daily contacts decreased by 86-89% (Zhang et al., 2020) while Rothwell (2020) suggests that in the USA the reduction lies between 75 and 90%. These should result in a four to 10 fold reduction in R, without taking into account any additional effects due to social distancing or improved personal hygiene. However, gathering information on average numbers of contacts per individual may not capture a key element of transmission dynamics, which is that a small proportion of cases may lead to the infection of very large numbers of others as a result of them interacting with large numbers of others in areas of high population density (Rader et al., 2020), mass gatherings (Che Mat, Edinur, Abdul Razab, & Safuan, 2020), confined settings such as homeless shelters (Baggett, Keyes, Sporn, & Gaeta, 2020) and other “superspreading” events (Liu, Eggo, & Kucharski, 2020; Wong, Jamaludin, Alikhan, & Chaw). In addition, those who acquire COVID-19 infection may have networks of social interaction that differ from those in a representative sample of the whole population.

### A simple approach to overcome the fundamental flaws of SEIR and other compartment models

SEIR and similar compartment models dominate the academic literature modelling the dynamics of the COVID-19 epidemic (Berger, Herkenhoff, & Mongey, 2020; Carcione, 2020; Chong, 2020; Danon, Brooks-Pollock, Bailey, & Keeling, 2020; Fang, Nie, & Penny, 2020; Giordano et al., 2020; Huang et al., 2020; Khan, Umar, & Khalid, 2020; Kucharski et al.; Pan et al., 2020; Kiesha Prem et al., 2020; Read, Bridgen, Cummings, Ho, & Jewell, 2020; Romero-Severson, Hengartner, Meadors, & Ke, 2020; Salomon, 2020; Tang et al., 2020; Teslya et al., 2020; J. T. Wu, Leung, & Leung, 2020; C. Yang & Wang, 2020; Z. Yang et al., 2020). These models are straightforward to use, and can be readily parameterised using estimates of transmission rate and mean times to becoming symptomatic and recovering (or more correctly, mean time to becoming infective and mean duration of the infective period – see below for a more detailed discussion). However, as we have shown elsewhere (Grant, 2020), compartment models do not simulate realistic distributions of the times that individuals spend in each compartment. The most common time taken to incubate the disease or recover after showing symptoms is zero in a continuous time model or one time step in a discrete time model, but some individuals spend very much longer than the mean residence time in individual compartments. As a result, the models make poor predictions of the dynamics of an epidemic (Krylova & Earn, 2013; Lloyd, 2001a, 2001b; Wearing, Rohani, & Keeling, 2005). They under-estimate the growth of numbers of cases and numbers of deaths during the initial exponential growth phase of an epidemic such as COVID-19, but after transmission rates are reduced by the introduction of social distancing or a sufficient proportion of the population has had the disease to see infection rates reduced by “herd immunity”, they over-estimate case numbers and the duration of epidemics. This also obscures the relationship between R and numbers of cases or numbers of deaths that we outline in the next section. Compartment models are, therefore, likely to be misleading if used to guide, or judge the effectiveness of, policy interventions to reduce the effects of an epidemic.

Time-since-infection models have some major theoretical advantages (see, e.g., Fraser, 2007; Katriel, 2013) but require the use of more challenging mathematical methods than many of the published applications of compartment models. We have shown that these shortfalls can be readily addressed by using a matrix model which is similar to a time since infection model (c.f. Katriel, 2013), but allows separate specification of time to becoming symptomatic and subsequent time to death or recovery and can be analysed using straightforward mathematical methods (Grant, 2020). In this paper we assume that the proportion of the population that is immune is negligible enabling us to use an even simpler version this model to predict the time course of the reduction in the epidemic in response to the introduction of restrictions on interactions between individuals, often referred to as “social distancing” and “lockdown”.

### The model

As in our previous paper, rather than combining all individuals which are in each compartment, the number of individuals in each compartment, and how long they have been there, are tracked in a vector. If the proportion of the population that is immune is small (so that we are in a period of exponential growth or decline, at a rate determined only by R), then we can simplify this model and only track exposed and infective individuals:

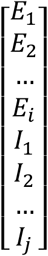

At each time step, this vector is multiplied by a projection matrix in which the number of new exposed individuals is determined by the number of infective individuals and the value of R. In the simplest case, where infectivity is constant across the I_j_ age classes, then each I individual gives rise to R/j individuals in class E1. Individuals that acquire infection move into the E_1_ compartment, then move through the remaining E and then I compartments. With fixed incubation and infection periods, all individuals move to the final E_i_ and I_j_ compartments before moving to the I_1_ and R compartments respectively, but varying incubation and infective periods can easily be accommodated (see Grant, 2020, for full details). If we assume a five day incubation period and an eight day infective period, then our population vector consists of five Exposed stages and eight Infective stages and the population projection matrix (with zeros other than those in the first row blanked out) is:

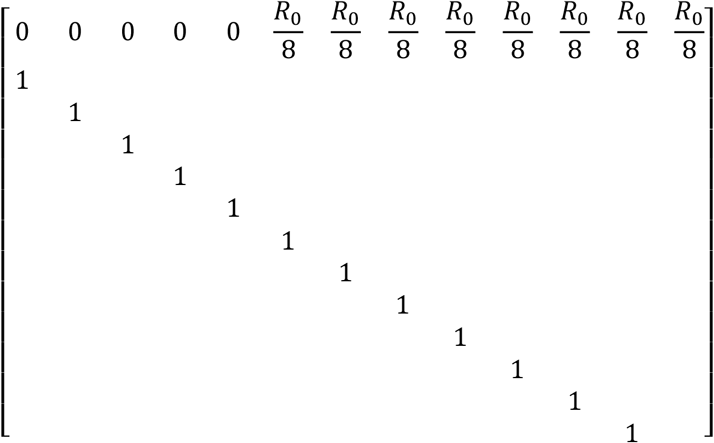

The daily rate of change in the number of exposed and infective individuals is given by the largest eigenvalue of this matrix, λ. In this case, the mean serial interval (or generation time) from one newly infected individual to the next is 9 days, so R_0_ ≈ λ^9.^ The equality is only exact when R_0_ = λ = 1, but the approximation is very close for R values in the range 0.25 to 3 (see blue line in Figure 1). The equivalent compartment model (including only the EI components of an SEIR model) has the projection matrix:

**Figure 1.**
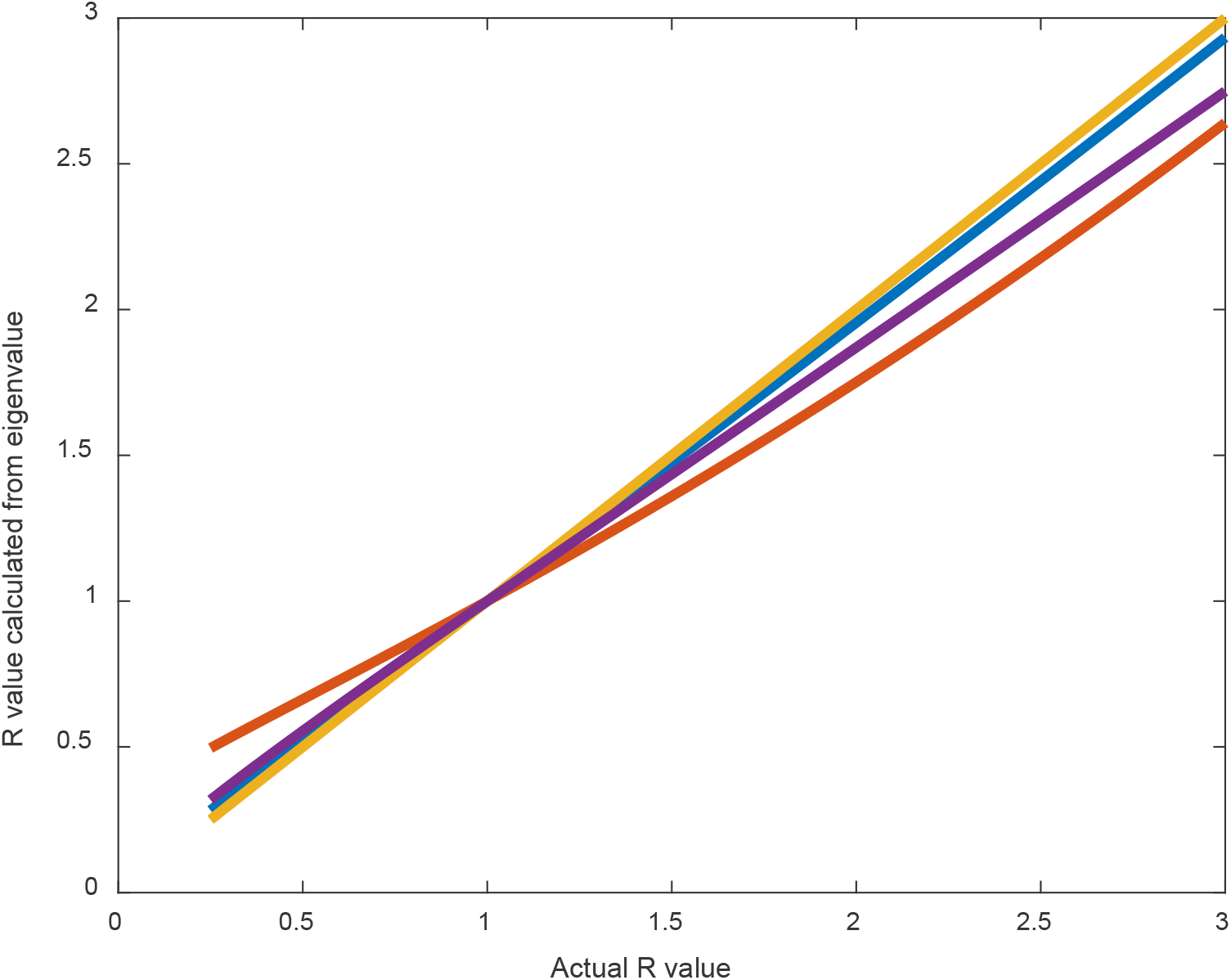
Values of R calculated from the dominant eigenvalue of a projection matrix plotted against true value. The blue line is for the projection matrix introduced here, the red indicates data for the corresponding SEIR model; The purple line uses the model formulation developed here with a gamma distributed incubation period with a mean of 5.93 days and a CV of 0.86; followed by an infectious period of 6 days. Yellow line is a reference line representing equality.

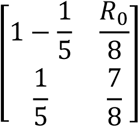

And again, R_0_ ≈ λ^9^. However, the approximation is poorer except for R_0_ = 1, particularly for values of R that are less than 1 (orange line in Figure 1). So as noted previously, compartment models underestimate infection rate increases when R is high, but overestimate R when an infection is under control and R is small. Variation in the incubation period reduces R at high values (as would be expected for any variance in vital rates, Ganyani et al., 2020; Lewontin & Cohen, 1969; Tuljapurkar, 1982), but has only a limited effect when R is less than 1.0 (purple line in Figure 1).

This gives us a simple route to estimate the value of R that is implied by a time series of infection rates, hospitalisation rates and death rates. The value of R is approximately equal to the daily change in any rate raised to the power of the mean generation time for the infection. Alternatively, if we have two rates Y_1_ and Y_2_ measured j days apart, then R can be estimated as:

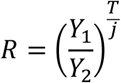

Where T is the mean generation time for the infection – the time elapsed between one individual being infected and the progeny cases being infected or the mean serial interval – the time elapsed between symptom onset in an infector-infectee pair. These two quantities may have different variances, but have the same mean (Britton & Tomba, 2019), so the distinction does not alter the analyses presented here. Estimates of the serial interval are in the range 3.95 (Ganyani et al., 2020) or 3.96 (Du et al., 2020) to 7.5 days (Li et al., 2020), but could be as high as 14.5 days if cases were equally infectious throughout a 15 day recovery period following an incubation period of 7 days (C. Yang & Wang, 2020). Flaxman *et al*., (2020) assume a mean serial interval of 6.5 days. Other estimates include a mean of 4.0 days but a median of 4.6 days based on the most reliable data (Nishiura, Linton, & Akhmetzhanov, 2020); 4.77 days, based on transmission to close contacts (Kwok, Wong, Wei, Wong, & Tang, 2020); 5.21 days in Singapore (Ganyani et al., 2020), 5.8 days and 6.5 days in Hong Kong (Chan et al., 2020; He et al., 2020). One of the challenges in estimating serial intervals is that it is difficult to unequivocally establish that two associated cases do indeed have the relationship of being a primary and secondary case. The lowest estimate above (3.96, from Du et al., 2020) was based on a data set that included negative serial intervals. Uncertainties about the value of the serial interval will affect the absolute value of R, but not comparisons over time; comparisons between geographical areas or between different data series. If R is equal to 1, the true value of the serial interval has no effect. For simplicity, if we assume that the serial interval is 7 days, then we can estimate R from time series that are reported at weekly intervals, using a 7 day running average for data that are reported daily. If the serial interval is less than this, then values of R that are less than 1 will be underestimated, while those greater than 1 will be overestimated. If, for example, the serial interval is actually 5 days, then assuming that it was 7 days would yield an estimate of 0.7 for a true value of 0.78 (= 0.7^5/7^) and a value of 4 for a true value of 2.69. But uncertainties about the serial interval will affect estimates of R_0_ from more detailed analyses in exactly the same way as they impact on our approximate calculation, as the serial interval or generation time is incorporated into most procedures to estimate R, either explicitly, or implicitly via the parameters of a compartment or other population model. The approach also assumes that a time series represents a constant proportion of numbers infected in the underlying population, but again, that challenge also affects more sophisticated analyses of data.

### Time to becoming symptomatic, testing positive and time to death

We cannot observe infection occurring. The data that we can observe are time series of numbers of individual becoming symptomatic; testing positive; being hospitalised or dying. As a first approximation, these data will lag behind changes in infection rates at an interval equal to the mean time from infection until that event occurs. Flaxman *et al*. (2020) use an incubation period which is gamma distributed with a mean of 5.1 days and a CV of 0.86. Time from appearance of symptoms to death is also assumed to be gamma distributed with a mean of 18.8 days and a CV of 0.45. Linton *et al*., (2020) give a similar value for the mean incubation period (5.0 - 6.0 days depending on model fitted and whether or not Wuhan residents are excluded from calculations). They estimate mean times from symptom onset to death as 14.5-15.1 days depending on model fitted. A gamma distribution gives a mean of 15.0 and SD 6.9. More recent data from Wuhan (Leung, Wu, Liu, & Leung, 2020) indicate a mean time from symptoms to death of 15.87 days (SD 7.17). Based on all these estimates, we would expect numbers developing symptoms to respond to lockdown with a time lag of between 5 and 7 days, with positive test results taking a few days longer as not all cases will present for testing immediately after symptom onset. Death rates should lag changes in infection rate by between 20 and 24 days.

### Using the tool on example time series

This approach allows the straightforward estimation of epidemic growth or decline rates from any time series of infection or death rates. The method involves some approximations, but the errors introduced by these are small when epidemics are declining and are smaller than those that result from uncertainties about the value of the serial interval, which also affect more sophisticated methods of estimating R. Data on COVID-19 infection and mortality rates vary in quality between countries depending upon intensity of testing; the criteria used to attribute deaths to COVID-19 and whether or not deaths in care homes are reported in the main national time series. The capacity for testing may have increased over time, leading to an increase in the number of cases which are identified by testing. The criteria used to select who is tested may have changed leading to a reduction in the proportion of tests that give a positive result. These are substantial challenges, but we do not examine them in detail here. We obtained the following data sets:

1. Daily data on deaths in hospital in England of patients with a confirmed COVID-19 test up until 17^th^ June (NHS England, 2020), from which data from 13^th^ June onwards were excluded from analysis as figures from the most recent few days are incomplete. Data were averaged weekly and the ratio of the average daily deaths in one week to that in the week before calculated. Data were analysed for the whole of England, and for each of seven English regions.
2. Deaths registered each week in England and Wales up until 5^th^ June 2020 where COVID-19 was identified as a cause of death (ONS, 2020b). Ratios of deaths in one week to those in the week before were calculated.
3. Daily numbers of positive test results in England from “Pillar 1” of the English testing program. These are based on hospital based testing of health and care workers and those with a “clinical need” but exclude swabs collected in the community and processed via commercial laboratories (Burn-Murdoch, Neville, Hughes, & Bounds, 2020; Public Health England, 2020a). Ratios between seven day averages were calculated.

In the presentation of these analyses, the ratio of numbers one week apart is referred to as a “weekly change”. If the serial interval is close to 7 days, then these numbers will be close to the value of R. If, as is likely, the serial interval is less than seven days, these numbers will be biased away from 1, with the magnitude of this bias being greater for values substantially greater than 1 (see above). For comparisons between adjacent weeks, the date used to identify a weekly change is that of the last day in the second week used to calculate the ratio, so changes reflect events occurring, on average, 3.5 days earlier than this In late March, death rates in hospital in England for those with a positive COVID-19 test result initially increased by a factor of 6.7 per week (Figure 2; equivalent to an R value of 3.9 if the serial interval is actually equal to 5 days), but then decreased, with the rate of change falling below 1 for the comparison between week ending 10^th^ April and week ending 17^th^ April. Regional data show a similar pattern (Fig. 3), with all but North West England having a rate of change lower than 1.0 at the same point in time. Weekly rates of change were then mostly in the range 0.6 to 0.9, with the regional geometrical means for the period 10^th^ April to 12^th^ July ranging from 0.64 in London to 0.93 in the North West, with an overall value for England of 0.75.

**Figure 2.**
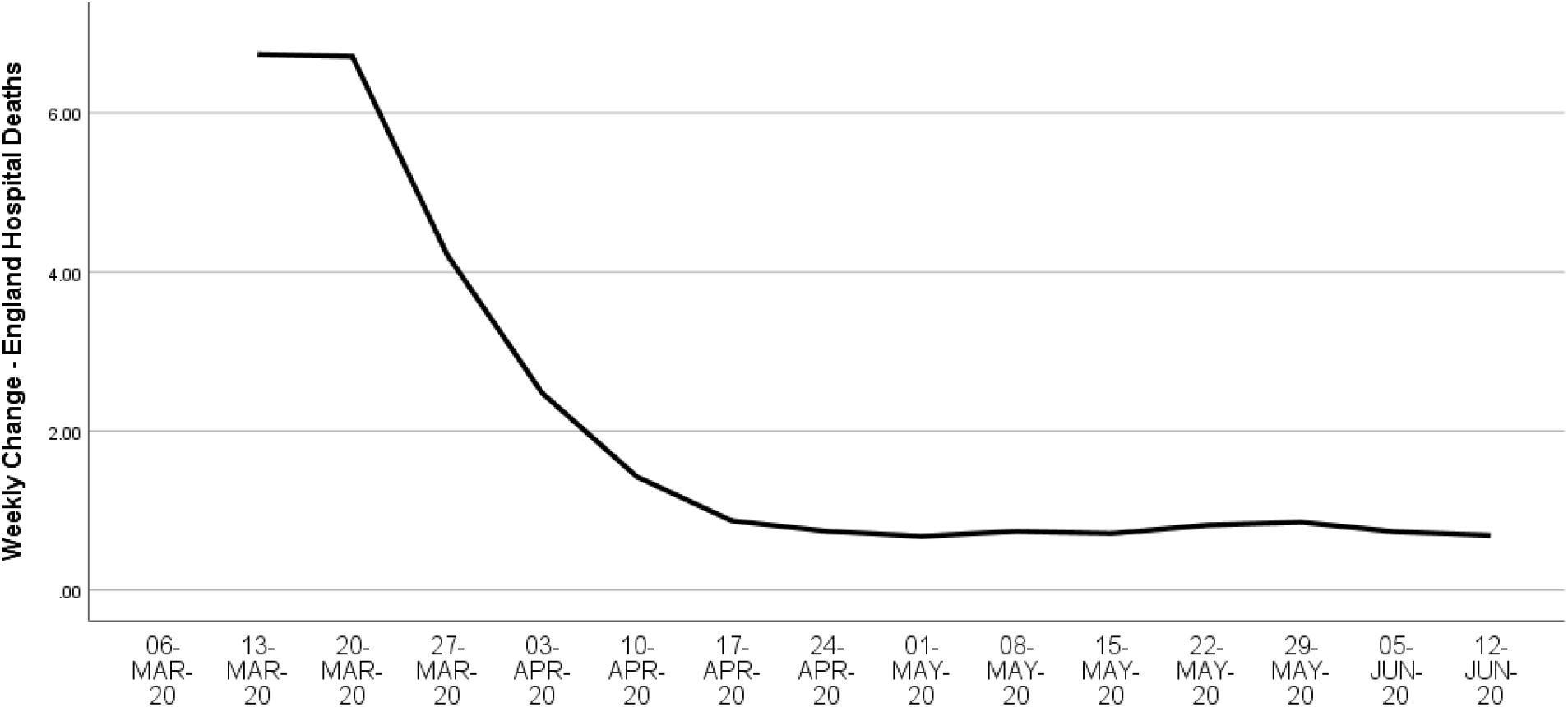
Weekly change in hospital deaths in England, where patient had a positive COVID-19 diagnosis, from the early stage of the epidemic in mid-March 2020 until 12^th^ June 2020. Values are plotted using the date at the end of the second week used to calculate the ratio.

**Figure 3.**
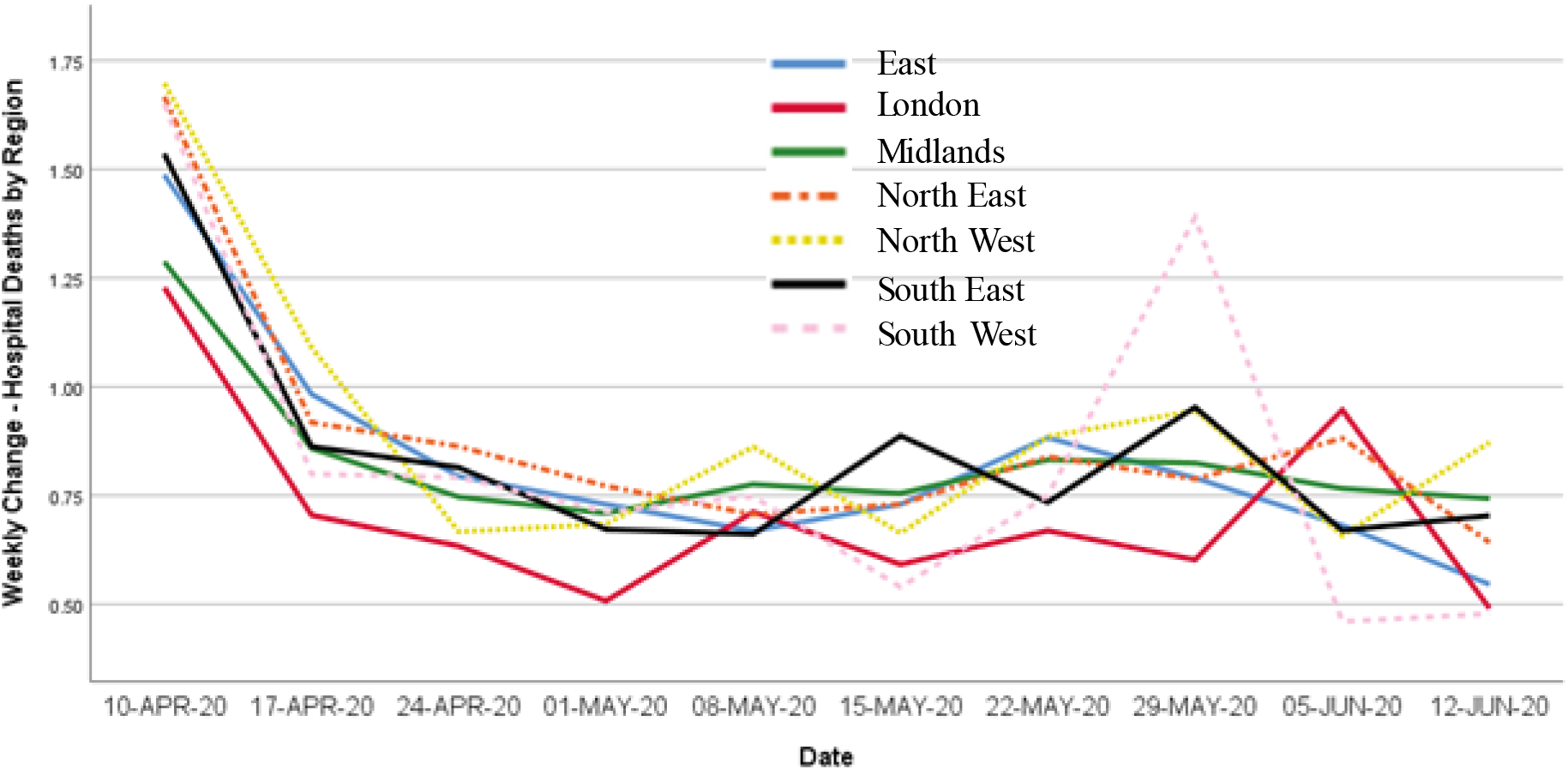
Weekly change in hospital deaths in seven English regions, where patient had a positive COVID-19 diagnosis. Data series are plotted from 10th April 2020 until 12^th^ June 2020, to display detail during period of epidemic decline.

Death registration data show a similar pattern (Figure 4), with initially high values, followed by a decline. The rate of change was 1.01 when deaths in week ending 17^th^ April are compared with the previous week, and rates below 1.0 thereafter. All regions showed a similar pattern (Figure 5), with London showing a slightly earlier decline. Calculating a geometric mean across the whole period from 24^th^ April to 5^th^ June gives an R value of 0.75 for England and Wales as a whole. The lowest average R over this whole period was for London, at 0.66, while values for the other regions were all in the range 0.72 to 0.80

**Figure 4.**
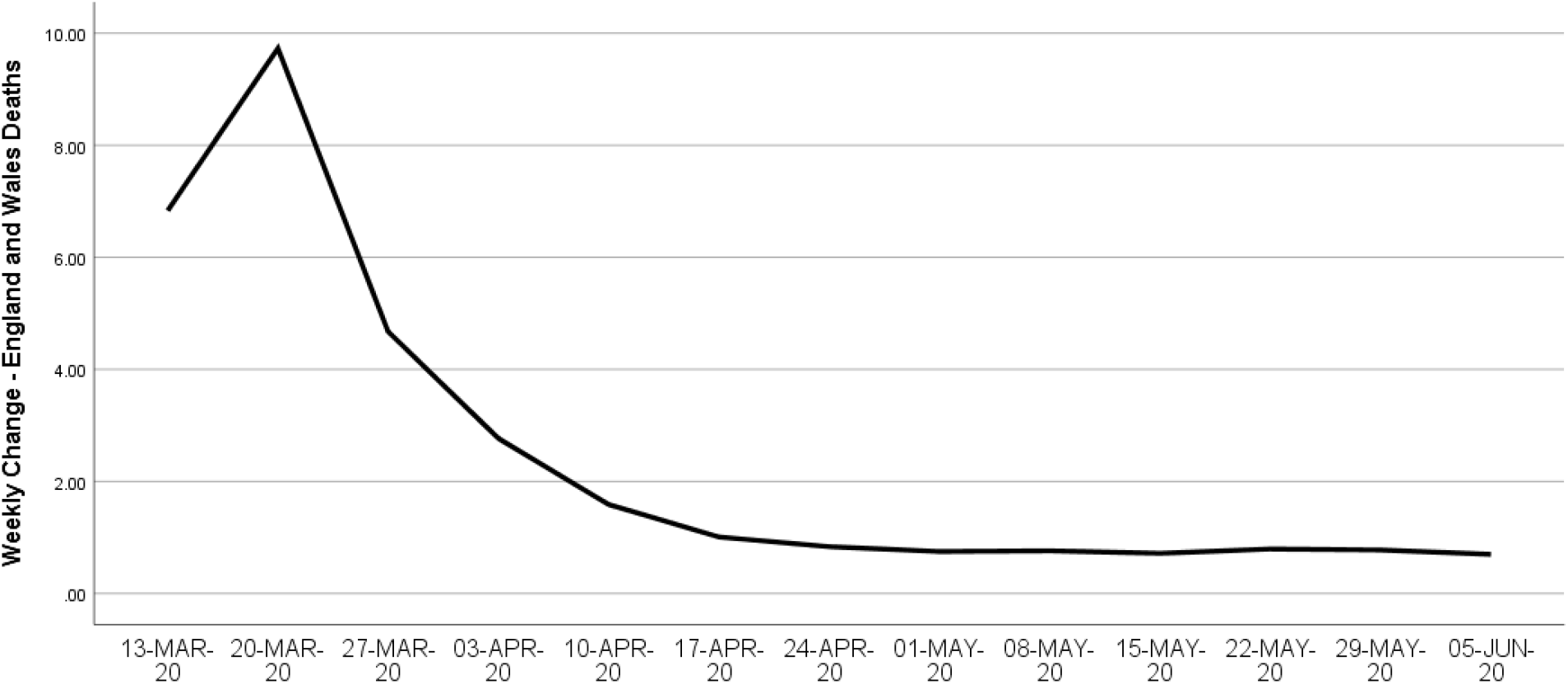
Weekly change in deaths registered in England and Wales, where COVID-19 was mentioned as a cause of death, from the early stage of the epidemic in mid-March 2020 until 12^th^ June 2020.

**Figure 5.**
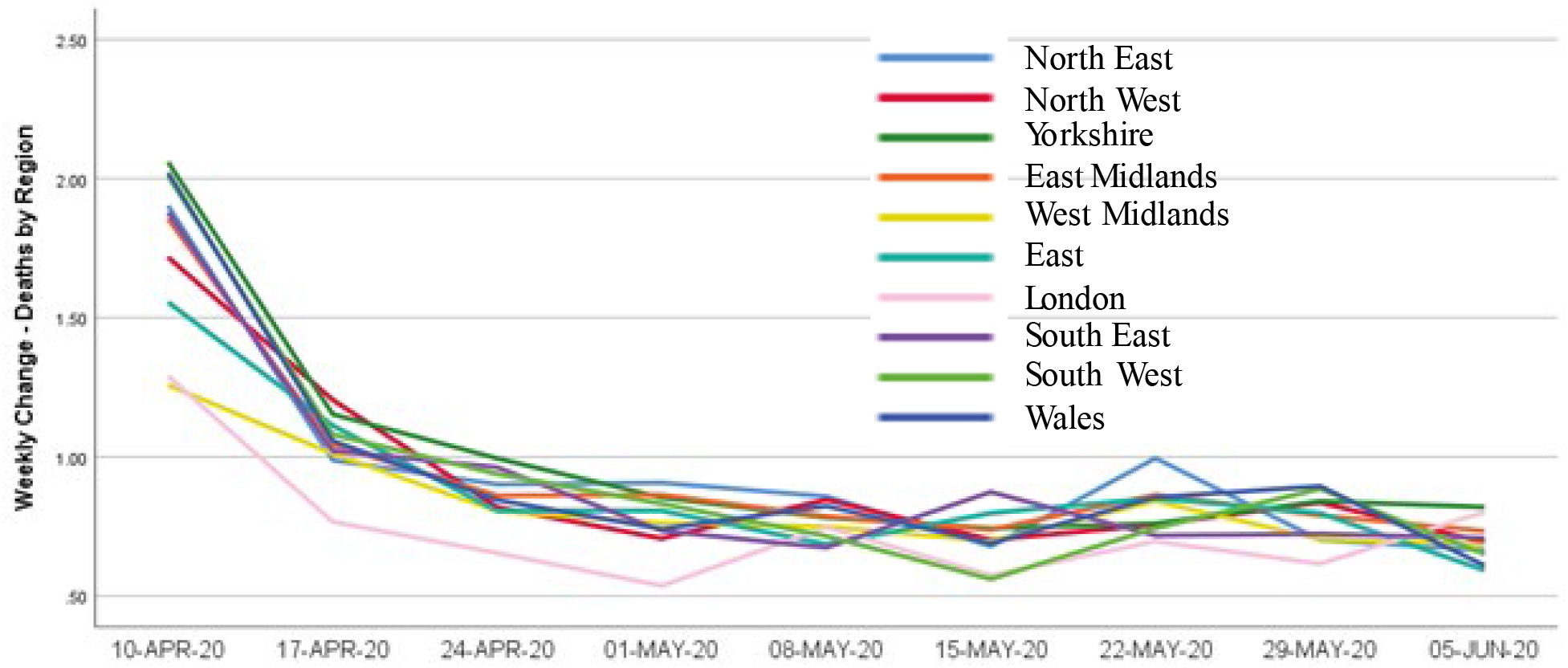
Weekly change in deaths registered in England and Wales where COVID-19 was mentioned as a cause of death, from 10^th^ April 2020 until 12^th^ June 2020. Data broken down by region (using a more detailed set of regions than figure 3).

Availability of COVID testing data for England is problematic. As at 1^st^ July 2020, only “pillar 1” data, which includes health care works and patients presenting to hospitals are made publically available, even though the great majority of positive test results now come from commercial “pillar 2” testing (Burn-Murdoch et al., 2020; Public Health England, 2020c). The rate of increase in the numbers of positive test results peaked in the week ending 6^th^ March (Figure 6). It then declined, with the rate of increase being 1.1 in week ending 10^th^ April and between 0.60 and 0.85 subsequently, with a geometric mean of 0.73 over the period 17^th^ April to 12^th^ June 2020.

**Figure 6.**
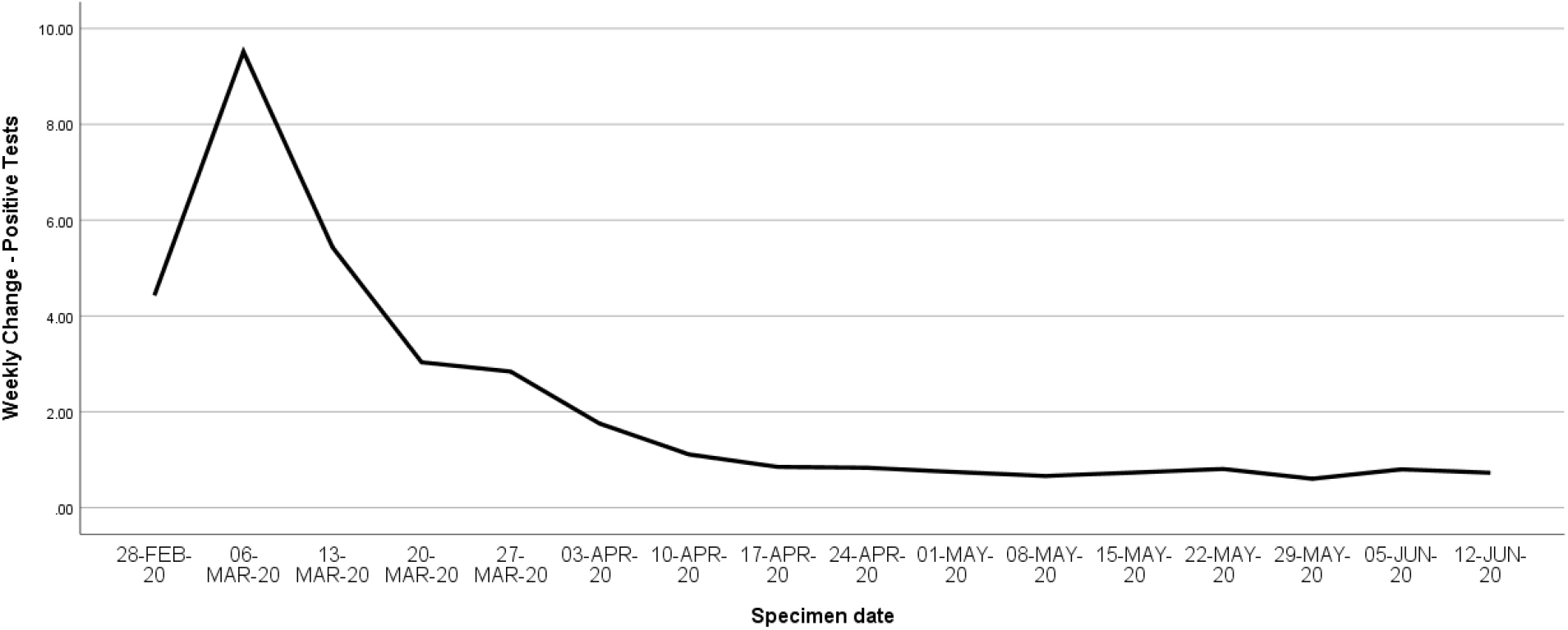
Weekly change in number of positive COVID-19 tests in England (data on “Pillar 1 tests – see text for details)

## Discussion

These three data sets give a consistent pattern, with peak increases in numbers of positive tests in early March, followed by peak increases in death rates in hospital a week later and peak increases in registered deaths a week after that. Rates of increase at the start of the epidemic were very high, corresponding to R values of 3.8 to 5 if the serial interval is equal to five days. It is possible that these initial values represent an over-estimate, as there may have been lower identification rates of symptomatic COVID cases in the early period of the epidemic, and limitations on testing capacity may have reduced the proportion of infected individuals being tested. So some of this initial high rate of increase may reflect an increase in diagnosis rates over time rather than an increase in disease prevalence. In all cases the rates of increase reduce below 1.0 around week ending 17^th^ April 2020. At a national level all show geometric mean weekly rates of change close to 0.75 over this period (equivalent to R = 0.81 for a serial interval of 5 days), although there is some spatial and temporal variability in rates of decline.

Is the pattern of these changes consistent with the expected consequences for R of policy interventions? On 16^th^ March 2020, the English government introduced recommendations on “social distancing” and voluntary self-isolation of those with pre-exiting health conditions that made them more vulnerable to COVID-19 (Public Health England, 2020b). Advice on personal hygiene and maintaining vigilance for COVID symptoms was in place before this and there is evidence that travel, and particularly use of public transport, began to decrease around 9^th^ March 2020 (Rieger, 2020). This was followed on 23^rd^ of March 2020 by mandatory restrictions on movement, meetings between more than two people, and non-essential economic activity, usually abbreviated as “lockdown” (Cabinet Office, 2020). Given the respective 7+ and 20+ day intervals between becoming infected and obtaining a positive test result or dying, then impacts of lockdown on the rate of increase of positive tests and deaths would be expected to occur from around 30^th^ March and 12^th^ April respectively. However, the rates of increase in the time series decline some time before these dates, with the discrepancy being particularly marked for the two death rate time series, which both move into decline only shortly before we would expect to see any effects of the introduction on lockdown. This suggests that the voluntary changes in behaviour, including increased personal hygiene and self-isolation of symptomatic individuals had a much greater impact on transmission rates than the subsequent lockdown.

Given the severity of the reduction in social and economic activity that has occurred in England as a result of lockdown (Cabinet Office, 2020; Jarvis et al., 2020) it is perhaps surprising that R values remain only a little less than 1. However, there is evidence that population averages of reductions in interactions between individuals may give a poor guide to transmission dynamics to and from those who are acquiring the infection. Confirmed cases who gave details to the English “Test and Trace” system between 28^th^ May and 10^th^ June 2020 reported an average of 9.5 “close” contacts during the period from 48 hours before symptoms appeared until 7 days afterwards once missing data are removed (Department of Health and Social Care, 2020). Here, “close” contact was defined as being in the same household; spending more than 15 minutes within 2m; a face to face conversation at closer than 1m, or sexual contact. This is a much more restrictive definition of contact than that used by Jarvis et al (2020), who counted all interactions in which “a few words were exchanged” or there was skin to skin contact. All the forms of “close” contact identified by Test and Trace were at the time unlawful except in an individual’s household or in a work environment where tight infection control measures were mandated. Average household size in the UK is 2.4 persons (ONS, 2017) so these interactions must be dominated by workplace and/or illicit interactions, unless many of the cases are occurring in overcrowded housing or large residential contexts. COVID-19 Attack rates within households may be as high as 17%, and overall attack rates with close contacts of 9.7% and 12.7% have been reported (Bi et al., 2020; Jing et al.; Mizumoto, Omori, & Nishiura, 2020). Multiplying these latter two attack rates by the numbers of reported close contacts from Test and Trace would yield R values of 0.92 and 1.2 respectively. This suggests that the failure of R to reduce substantially below 1.0 may result from transmission of the infection between individuals who continue to have relatively high frequency of close contacts with others. Individuals with higher than average numbers of close contacts are likely to interact disproportionately with each other, as a result of overcrowded housing, work places like meat packing facilities and residential care homes where social distancing is difficult (O’Neill et al.; Reuben, 2020) or as a result of lifestyle choices. If this is the case, unselective nationwide “lockdowns” will have much less impact on the time course of an epidemic. Many of their restrictions will fall on interactions between individuals where attack rates are already very low as a result of social distancing, while they have much less impact on those interactions where attack rates are much higher. There appears to be considerable heterogeneity of infection rates - newly released data on Upper Tier Local Authorities show weekly numbers of positive COVID-19 tests per 100 000 population varying between zero and 140 (UK Statistics Authority, 2020). Policies will be more cost-effective if efforts can be focussed on identifying and intervening in local areas, work places and social groupings where infection rates are relatively high while relying on voluntary social distancing to control transmission in areas where there are relatively few cases and social contexts where attack rates are low.

## Data Availability

All data are from public databases, available at locations cited in the paper

## Bibliography

1. Baggett, T. P., Keyes, H., Sporn, N., & Gaeta, J. M. (2020). Prevalence of SARS-CoV-2 Infection in Residents of a Large Homeless Shelter in Boston. JAMA, 323(21), 2191–2192. doi:10.1001/jama.2020.6887

2. Belfin, R. V., Brodka, P., Radhakrishnan, B., & Rejula, V. (2020). COVID-19 peak estimation and effect of nationwide lockdown in India. medRxiv, 2020.2005.2009.20095919. doi:10.1101/2020.05.09.20095919

3. Berger, D., Herkenhoff, K., & Mongey, S. (2020). An SEIR Infectious Disease Model with Testing and Conditional Quarantine, University of Chicago, Becker Friedman Institute for Economics Working Paper No. 2020–25.

4. Bi, Q., Wu, Y., Mei, S., Ye, C., Zou, X., Zhang, Z., … Feng, T. (2020). Epidemiology and transmission of COVID-19 in 391 cases and 1286 of their close contacts in Shenzhen, China: a retrospective cohort study. The Lancet Infectious Diseases. doi:10.1016/S1473-3099(20)30287-5

5. Birrell, P., Blake, J., Leeuwen, E. v., Joint PHE Modelling Cell, MRC Biostatistics Unit COVID-19 Working Group, & Angelis, D. D. (2020). COVID-19: nowcast and forecast. Retrieved from https://www.mrc-bsu.cam.ac.uk/now-casting/

6. Britton, T., & Tomba, G. S. (2019). Estimation in emerging epidemics: biases and remedies. Journal of the Royal Society Interface, 16(150). doi:10.1098/rsif.2018.0670

7. Burn-Murdoch, J., Neville, S., Hughes, L., & Bounds, A. (2020). Lack of local Covid-19 testing data hinders UK’s outbreak response. Retrieved from https://www.ft.com/content/301c847c-a317-4950-a75b-8e66933d423a

8. Cabinet Office. (2020). Staying at home and away from others (social distancing). Retrieved from https://www.gov.uk/government/publications/full-guidance-on-staying-at-home-and-away-from-others/full-guidance-on-staying-at-home-and-away-from-others

9. Carcione, J. M. (2020). A simulation of a COVID-19 epidemic based on a deterministic SEIR model. medRxiv, 2020.2004.2020.20072272. doi:10.1101/2020.04.20.20072272

10. Chan, Y., Flasche, S., Lam, T., Leung, M., Wong, M., Lam, H., & Chuang, S. (2020). Transmission dynamics, serial interval and epidemiology of COVID-19 diseases in Hong Kong under different control measures [version 1; peer review: awaiting peer review]. Wellcome Open Research, 5(91). doi:10.12688/wellcomeopenres.15896.1

11. Che Mat, N. F., Edinur, H. A., Abdul Razab, M. K. A., & Safuan, S. (2020). A single mass gathering resulted in massive transmission of COVID-19 infections in Malaysia with further international spread. J Travel Med, 27(3). doi:10.1093/jtm/taaa059

12. Chong, Y. C. (2020). A Novel Method for the Estimation of a Dynamic Effective Reproduction Number (Dynamic-R) in the CoViD-19 Outbreak. medRxiv, 2020.2002.2022.20023267. doi:10.1101/2020.02.22.20023267

13. Cori, A., Ferguson, N. M., Fraser, C., & Cauchemez, S. (2013). A New Framework and Software to Estimate Time-Varying Reproduction Numbers During Epidemics. American Journal of Epidemiology, 178(9), 1505–1512. doi:10.1093/aje/kwt133

14. Danon, L., Brooks-Pollock, E., Bailey, M., & Keeling, M. J. (2020). A spatial model of CoVID-19 transmission in England and Wales: early spread and peak timing. medRxiv, 2020.2002.2012.20022566. doi:10.1101/2020.02.12.20022566

15. Department of Health and Social Care. (2020). Experimental statistics: Weekly NHS Test and Trace bulletin, England: 28 May – 10 June 2020. Retrieved from

16. Du, Z., Xu, X., Wu, Y., Wang, L., Cowling, B. J., & Meyers, L. A. (2020). Serial Interval of COVID-19 among Publicly Reported Confirmed Cases. Emerg Infect Dis, 26(6), 1341–1343. doi:10.3201/eid2606.200357

17. Fang, Y., Nie, Y., & Penny, M. (2020). Transmission dynamics of the COVID-19 outbreak and effectiveness of government interventions: A data-driven analysis. Journal of Medical Virology, n/a(n/a). doi:10.1002/jmv.25750

18. Flaxman, S., Mishra, S., Gandy, A., Unwin, H., Coupland, H., Mellan, T., … Bhatt, S. (2020). Estimating the number of infections and the impact of non-pharmaceutical interventions on COVID-19 in 11 European countries. Imperial College COVID-19 Response Team.

19. Fraser, C. (2007). Estimating Individual and Household Reproduction Numbers in an Emerging Epidemic. Plos One, 2(8). doi:10.1371/journal.pone.0000758

20. Ganyani, T., Kremer, C., Chen, D., Torneri, A., Faes, C., Wallinga, J., & Hens, N. (2020). Estimating the generation interval for coronavirus disease (COVID-19) based on symptom onset data, March 2020. Eurosurveillance, 25(17), 2000257. doi: doi:https://doi.org/10.2807/1560-7917.ES.2020.25.17.2000257

21. Giordano, G., Blanchini, F., Bruno, R., Colaneri, P., Di Filippo, A., Di Matteo, A., & Colaneri, M. (2020). Modelling the COVID-19 epidemic and implementation of population-wide interventions in Italy. Nature Medicine. doi:10.1038/s41591-020-0883-7

22. Government Office for Science. (2020). The latest reproduction number (R) and growth rate of coronavirus (COVID-19) in the UK. Retrieved from https://www.gov.uk/guidance/the-r-number-in-the-uk

23. Grant, A. (2020). Dynamics of COVID-19 epidemics: SEIR models underestimate peak infection rates and overestimate epidemic duration. medRxiv, 2020.2004.2002.20050674. doi:10.1101/2020.04.02.20050674

24. He, X., Lau, E. H. Y., Wu, P., Deng, X. L., Wang, J., Hao, X. X., … Leung, G. M. (2020). Temporal dynamics in viral shedding and transmissibility of COVID-19. Nature Medicine. doi:10.1038/s41591-020-0869-5

25. Huang, Y., Yang, L., Dai, H., Tian, F., & Chen, K. (2020). Epidemic situation and forecasting of COVID-19 in and outside China. Bull World Health Organ. doi:http://dx.doi.org/10.2471/BLT.20.255158

26. Jarvis, C. I., Van Zandvoort, K., Gimma, A., Prem, K., Klepac, P., Rubin, G. J., … Grp, C. C.-W. (2020). Quantifying the impact of physical distance measures on the transmission of COVID-19 in the UK. Bmc Medicine, 18(1). doi:10.1186/s12916-020-01597-8

27. Jing, Q.-L., Liu, M.-J., Zhang, Z.-B., Fang, L.-Q., Yuan, J., Zhang, A.-R., … Yang, Y. Household secondary attack rate of COVID-19 and associated determinants in Guangzhou, China: a retrospective cohort study. The Lancet Infectious Diseases. doi:10.1016/S1473-3099(20)30471-0

28. Katriel, G. (2013). Stochastic discrete-time age-of-infection epidemic models. International Journal of Biomathematics, 06. doi:10.1142/S1793524512500660

29. Khan, E. A., Umar, M., & Khalid, M. (2020). COVID 19: An SEIR model predicting disease progression and healthcare outcomes for Pakistan. medRxiv, 2020.2005.2029.20116517. doi:10.1101/2020.05.29.20116517

30. Krylova, O., & Earn, D. J. D. (2013). Effects of the infectious period distribution on predicted transitions in childhood disease dynamics. Journal of the Royal Society Interface, 10(84). doi:10.1098/rsif.2013.0098

31. Kucharski, A. J., Russell, T. W., Diamond, C., Liu, Y., Edmunds, J., Funk, S., … Flasche, S. Early dynamics of transmission and control of COVID-19: a mathematical modelling study. The Lancet Infectious Diseases. doi:10.1016/S1473-3099(20)30144-4

32. Kwok, K. O., Wong, V. W. Y., Wei, W. I., Wong, S. Y. S., & Tang, J. W. T. (2020). Epidemiological characteristics of the first 53 laboratory-confirmed cases of COVID-19 epidemic in Hong Kong, 13 February 2020. Eurosurveillance, 25(16), 23–31. doi:10.2807/1560-7917.es.2020.25.16.2000155

33. Lauer, S. A., Grantz, K. H., Bi, Q., Jones, F. K., Zheng, Q., Meredith, H. R., … Lessler, J. (2020). The Incubation Period of Coronavirus Disease 2019 (COVID-19) From Publicly Reported Confirmed Cases: Estimation and Application. Annals of Internal Medicine. doi:10.7326/m20-0504

34. Leung, K., Wu, J. T., Liu, D., & Leung, G. M. (2020). First-wave COVID-19 transmissibility and severity in China outside Hubei after control measures, and second-wave scenario planning: a modelling impact assessment. The Lancet. doi:10.1016/S0140-6736(20)30746-7

35. Lewontin, R. C., & Cohen, D. (1969). On population growth in a randomly varying environment. Proceedings of the National Academy of Sciences of the United States of America, 62(4), 1056-&. doi:10.1073/pnas.62.4.1056

36. Li, Q., Guan, X., Wu, P., Wang, X., Zhou, L., Tong, Y., … Feng, Z. (2020). Early Transmission Dynamics in Wuhan, China, of Novel Coronavirus–Infected Pneumonia. New England Journal of Medicine, 382(13), 1199–1207. doi:10.1056/NEJMoa2001316

37. Linton, N. M., Kobayashi, T., Yang, Y. C., Hayashi, K., Akhmetzhanov, A. R., Jung, S. M., … Nishiura, H. (2020). Incubation Period and Other Epidemiological Characteristics of 2019 Novel Coronavirus Infections with Right Truncation: A Statistical Analysis of Publicly Available Case Data. Journal of Clinical Medicine, 9(2). doi:10.3390/jcm9020538

38. Liu, Y., Eggo, R. M., & Kucharski, A. J. (2020). Secondary attack rate and superspreading events for SARS-CoV-2. Lancet, 395(10227), E47–E47. doi:10.1016/s0140-6736(20)30462-1

39. Lloyd, A. L. (2001a). Destabilization of epidemic models with the inclusion of realistic distributions of infectious periods. Proceedings of the Royal Society B-Biological Sciences, 268(1470), 985–993. doi:10.1098/rspb.2001.1599

40. Lloyd, A. L. (2001b). Realistic distributions of infectious periods in epidemic models: Changing patterns of persistence and dynamics. Theoretical Population Biology, 60(1), 59–71. doi:10.1006/tpbi.2001.1525

41. Mizumoto, K., Omori, R., & Nishiura, H. (2020). Age specificity of cases and attack rate of novel coronavirus disease (COVID-19). medRxiv, 2020.2003.2009.20033142. doi:10.1101/2020.03.09.20033142

42. Mossong, J., Hens, N., Jit, M., Beutels, P., Auranen, K., Mikolajczyk, R., … Edmunds, W. J. (2008). Social contacts and mixing patterns relevant to the spread of infectious diseases. Plos Medicine, 5(3), e74–e74. doi:10.1371/journal.pmed.0050074

43. NHS England. (2020). COVID-19 Daily Deaths. Retrieved from https://www.england.nhs.uk/statistics/statistical-work-areas/covid-19-daily-deaths/

44. Nishiura, H., Linton, N. M., & Akhmetzhanov, A. R. (2020). Serial interval of novel coronavirus (COVID-19) infections. Int J Infect Dis, 93, 284–286. doi:10.1016/j.ijid.2020.02.060

45. O’Neill, D., Briggs, R., Holmerova, I., Samuelsson, O., Gordon, A. L., Martin, F. C., & European Geriatric Med, S. COVID-19 highlights the need for universal adoption of standards of medical care for physicians in nursing homes in Europe. European Geriatric Medicine. doi:10.1007/s41999-020-00347-6

46. ONS. (2017). Families and households in the UK: 2017. Retrieved from

47. ONS. (2020a). Coronavirus (COVID-19) related deaths by occupation, England and Wales: deaths registered up to and including 20 April 2020. Retrieved from

48. ONS. (2020b). Deaths registered weekly in England and Wales, provisional: week ending 5 June 2020. Retrieved from

49. Pan, Y., Yao, Y., Liu, Z., Li, M., Wang, Y., Dong, W., … Wang, W. (2020). Effectiveness of control strategies for Coronavirus Disease 2019: a SEIR dynamic modeling study. Bull World Health Organ. doi:http://dx.doi.org/10.2471/BLT.20.253807

50. Prem, K., Liu, Y., Russell, T. W., Kucharski, A. J., Eggo, R. M., Davies, N., … Klepac, P. (2020). The effect of control strategies to reduce social mixing on outcomes of the COVID-19 epidemic in Wuhan, China: a modelling study. The Lancet Public Health. doi:10.1016/S2468-2667(20)30073-6

51. Prem, K., Liu, Y., Russell, T. W., Kucharski, A. J., Eggo, R. M., Davies, N., … Ctr Math Modelling Infect Dis, C. (2020). The effect of control strategies to reduce social mixing on outcomes of the COVID-19 epidemic in Wuhan, China: a modelling study. Lancet Public Health, 5(5), E261–E270. doi:10.1016/s2468-2667(20)30073-6

52. Public Health England. (2020a). Coronavirus (COVID-19) in the UK. Retrieved from https://coronavirus.data.gov.uk/

53. Public Health England. (2020b). Guidance on social distancing for everyone in the UK. Retrieved from https://www.gov.uk/government/publications/covid-19-guidance-on-social-distancing-and-for-vulnerable-people/guidance-on-social-distancing-for-everyone-in-the-uk-and-protecting-older-people-and-vulnerable-adults

54. Public Health England. (2020c). Weekly Coronavirus Disease 2019 (COVID-19) Surveillance Report.

55. Year: 2020, Week: 26. Retrieved from London: https://assets.publishing.service.gov.uk/government/uploads/system/uploads/attachment_data/file/895356/Weekly_COVID19_Surveillance_Report_w26.pdf

56. Rader, B., Scarpino, S., Nande, A., Hill, A., Reiner, R., Pigott, D., … Kraemer, M. U. G. (2020). Crowding and the epidemic intensity of COVID-19 transmission. medRxiv, 2020.2004.2015.20064980. doi:10.1101/2020.04.15.20064980

57. Read, J. M., Bridgen, J. R., Cummings, D. A., Ho, A., & Jewell, C. P. (2020). Novel coronavirus 2019-nCoV: early estimation of epidemiological parameters and epidemic predictions. medRxiv, 2020.2001.2023.20018549. doi:10.1101/2020.01.23.20018549

58. Reuben, A. (2020). Coronavirus: Why have there been so many outbreaks in meat processing plants? Retrieved from https://www.bbc.co.uk/news/53137613

59. Ridenhour, B., Kowalik, J. M., & Shay, D. K. (2014). Unraveling R0: considerations for public health applications. American journal of public health, 104(2), e32–e41. doi:10.2105/AJPH.2013.301704

60. Rieger, M. O. (2020). A secret erosion of the lockdown? The activity patterns of Britons in March and April 2020. Retrieved from https://blogs.lse.ac.uk/politicsandpolicy/activity-patterns-during-the-lockdown/

61. Rocklöv, J., Sjödin, H., & Wilder-Smith, A. (2020). COVID-19 outbreak on the Diamond Princess cruise ship: estimating the epidemic potential and effectiveness of public health countermeasures. J Travel Med, 27(3). doi:10.1093/jtm/taaa030

62. Romero-Severson, E. O., Hengartner, N., Meadors, G., & Ke, R. (2020). Decline in global COVID-19 transmission. medRxiv, 2020.2004.2018.20070771. doi:10.1101/2020.04.18.20070771

63. Rothwell, J. (2020). Americans’ Social Contacts During the COVID-19 Pandemic. Retrieved from https://news.gallup.com/opinion/gallup/308444/americans-social-contacts-during-covid-pandemic.aspx

64. Salomon, J. A. (2020). Defining high-value information for COVID-19 decision-making. medRxiv, 2020.2004.2006.20052506. doi:10.1101/2020.04.06.20052506

65. Tang, B., Wang, X., Li, Q., Bragazzi, N. L., Tang, S. Y., Xiao, Y. N., & Wu, J. H. (2020). Estimation of the Transmission Risk of the 2019-nCoV and Its Implication for Public Health Interventions. Journal of Clinical Medicine, 9(2). doi:10.3390/jcm9020462

66. Teslya, A., Pham, T. M., Godijk, N. G., Kretzschmar, M. E., Bootsma, M. C. J., & Rozhnova, G. (2020). Impact of self-imposed prevention measures and short-term government intervention on mitigating and delaying a COVID-19 epidemic. medRxiv, 2020.2003.2012.20034827. doi:10.1101/2020.03.12.20034827

67. Tuljapurkar, S. D. (1982). Population-dynamics in variable environments .3. evolutionary dynamics of r-selection. Theoretical Population Biology, 21(1), 141–165. doi:10.1016/0040-5809(82)90010-7

68. UK Statistics Authority. (2020). COVID-19 Local Area Data. Retrieved from https://www.statisticsauthority.gov.uk/news/covid-19-local-area-data/

69. Wallinga, J., & Lipsitch, M. (2007). How generation intervals shape the relationship between growth rates and reproductive numbers. Proceedings of the Royal Society B-Biological Sciences, 274(1609), 599–604. doi:10.1098/rspb.2006.3754

70. Wearing, H. J., Rohani, P., & Keeling, M. J. (2005). Appropriate models for the management of infectious diseases. Plos Medicine, 2(7), 621–627. doi:10.1371/journal.pmed.0020174

71. Wong, J., Jamaludin, S. A., Alikhan, M. F., & Chaw, L. Asymptomatic transmission of SARS-CoV-2 and implications for mass gatherings. Influenza and Other Respiratory Viruses, n/a(n/a). doi:10.1111/irv.12767

72. Wu, J. H., Tang, B., Bragazzi, N. L., Nah, K., & McCarthy, Z. (2020). Quantifying the role of social distancing, personal protection and case detection in mitigating COVID-19 outbreak in Ontario, Canada. Journal of Mathematics in Industry, 10(1). doi:10.1186/s13362-020-00083-3

73. Wu, J. T., Leung, K., & Leung, G. M. (2020). Nowcasting and forecasting the potential domestic and international spread of the 2019-nCoV outbreak originating in Wuhan, China: a modelling study. Lancet, 395(10225), 689–697. doi:10.1016/s0140-6736(20)30260-9

74. Yang, C., & Wang, J. (2020). A mathematical model for the novel coronavirus epidemic in Wuhan, China. Mathematical Biosciences and Engineering, 17(3), 2708–2724. doi:http://dx.doi.org/10.3934/mbe.2020148

75. Yang, Z., Zeng, Z., Wang, K., Wong, S.-S., Liang, W., Zanin, M., … He, J. (2020). Modified SEIR and AI prediction of the epidemics trend of COVID-19 in China under public health interventions. Journal of Thoracic Disease, 12(3), 165–174.

76. Zhang, J., Litvinova, M., Liang, Y., Wang, Y., Wang, W., Zhao, S., … Yu, H. (2020). Age profile of susceptibility, mixing, and social distancing shape the dynamics of the novel coronavirus disease 2019 outbreak in China. medRxiv, 2020.2003.2019.20039107. doi:10.1101/2020.03.19.20039107

